# Motion prediction using electromyography and sonomyography for an individual with transhumeral limb loss

**DOI:** 10.1101/2020.12.23.20248489

**Authors:** Susannah Engdahl, Ananya Dhawan, György Lévay, Ahmed Bashatah, Rahul Kaliki, Siddhartha Sikdar

## Abstract

Controlling multi-articulated prosthetic hands with surface electromyography can be challenging for users. Sonomyography, or ultrasound-based sensing of muscle deformation, avoids some of the problems of electromyography and enables classification of multiple motion patterns in individuals with upper limb loss. Because sonomyography has been previously studied only in individuals with transradial limb loss, the purpose of this study was to assess the feasibility of an individual with transhumeral limb loss using this modality for motion classification. A secondary aim was to compare motion classification performance between electromyography and sonomyography. A single individual with transhumeral limb loss created two datasets containing 11 motions each (individual flexion of each finger, thumb abduction, power grasp, key grasp, tripod, point, pinch, wrist pronation). Electromyography or sonomyography signals associated with every motion were acquired and cross-validation accuracy was computed for each dataset. While all motions were usually predicted successfully with both electromyography and sonomyography, the cross-validation accuracies were typically higher for sonomyography. Although this was an exploratory study, the results suggest that controlling an upper limb prosthesis using sonomyography may be feasible for individuals with transhumeral limb loss.

## Introduction

Abandonment of upper limb prostheses is common [1] and is largely attributable to user concerns regarding comfort and function [2,3]. In fact, one survey reported that 98% of individuals who have rejected a prosthesis believe they are just as or more functional without a prosthesis, an opinion which is echoed by 60% of those who do wear a prosthesis frequently [2]. Recent research efforts have focused on developing dexterous prosthetic hands and associated control strategies in an effort to improve user functionality, but the prosthetic terminal devices and componentry have improved much more rapidly than the ability to control the devices. Thus, robustly inferring volitional motor intent from the user remains challenging [4].

Conventional prosthesis control strategies rely on surface electromyography (EMG) to detect muscle activation in a user’s residual limb. However, surface EMG signals typically have poor amplitude resolution and a low signal-to-noise ratio [5,6], as well as limited specificity between muscles due to cross-talk and co-activation [7–10]. Multi-articulated prosthetic hands commonly use dual-site surface EMG electrodes placed on an agonist-antagonist muscle pair in order to actuate the hand using a direct control strategy. Since direct control only enables opening and closing of the hand, switching between grasps must be initiated by a separate trigger such as a designated EMG signal pattern (e.g., co-contraction), physical gesture, or button press. These mode switching methods can be cumbersome and are strongly disliked by some users [11].

In an effort to mitigate some of these problems, pattern recognition algorithms are actively being developed in order to extract user intent from an array of surface EMG signals. Pattern recognition has shown some functional advantages in comparison to direct control [12,13]. However, one study found no difference in function or satisfaction between the systems for a two degree of freedom prosthesis [14] and others found that pattern recognition failed to improve outcomes for users of multi-degree of freedom prostheses [15,16]. These studies focused on gross wrist and hand movements, but dexterous control of individual finger movements is also possible with pattern recognition [17–20]. Despite these potential advantages of pattern recognition, considerable training is often required for users to reliably produce the set of distinct EMG patterns required for this modality [21,22].

Rather than controlling a prosthesis through detection of muscular electrical activity, an alternative approach relies on ultrasound imaging to sense muscle deformation during voluntary movement. This method, known as sonomyography (SMG), avoids many of the limitations of EMG because it can spatially resolve individual muscles, even those deep inside the arm. This high spatial specificity means that muscular cross-talk does not contaminate the extracted control signals that can be used to drive movement of a prosthesis. Numerous studies have established SMG as a viable option for gesture recognition and prosthesis control [23–26]. In particular, our group has demonstrated the ability to classify five individual digit movements with 97% accuracy [27] and 15 complex grasps with 91% accuracy [28] in able-bodied individuals. There has been relatively little exploration of the use of SMG in individuals with upper limb loss [29–33], although we have reported an average classification accuracy of 96% for five grasps in this population [32] with minimal training required [33].

However, all studies to date on individuals with upper limb loss using SMG have focused on a transradial population. It is unknown whether SMG can be successfully used for motion classification by individuals with more proximal limb loss. Since the muscles associated with wrist, hand, and finger control are primarily located in the forearm, absence of the forearm may preclude the use of SMG for classifying motion. Thus, the purpose of this exploratory study was to quantify the motion classification accuracy achievable by an individual with transhumeral limb loss using SMG. A secondary aim was to directly compare motion classification using EMG and SMG in order to understand the relative merits of each modality.

## Methods

### Subject

The subject underwent bilateral wrist disarticulation (right arm) and transhumeral (left arm) amputations nine years prior to this data collection. The subject reported use of a pattern recognition controlled prosthesis consisting of an externally-powered elbow, wrist and terminal device on a daily basis for over two years. Before that, the subject had used a direct controlled hybrid prosthesis (body-powered elbow, externally-powered wrist and terminal device) for six years. Both devices were attached to the transhumeral residual limb via a suspension socket. Although the subject had been exposed to SMG approximately one year prior to this data collection, he had only used it with his right arm and was naï ve to its use with the arm affected by transhumeral amputation. The study protocol was approved by the George Mason University Institutional Review Board and the subject provided written informed consent prior to participating.

### Data acquisition with sonomyography

All data were acquired from the subject’s transhumeral residual limb. A Terason uSmart 3200T ultrasound system (Terason, Burlington, MA) was used for the studies. A low-profile, high-frequency, linear 16HL7 ultrasound transducer was positioned on the subject’s residual limb using a stretchable fabric cuff. Ultrasound image sequences were acquired at a sampling rate of 13.44 +/- 0.66 Hz and transferred to a PC in real-time using an Epiphan screen capture card (Epiphan Systems, Palo Alto, CA), where they were processed in MATLAB (MathWorks, Natick, MA) using custom algorithms. Note that the sampling rate was variable due to limitations imposed by the integration of the screen capture card with MATLAB. However, this does not drastically impact our study as the classification procedure discussed herein does not rely on temporal information.

The subject created a single dataset containing a set of 11 motions (*index* finger flexion, *middle* finger flexion, *ring* finger flexion, *little* finger flexion, *thumb abduction, power grasp, key grasp, tripod, point, pinch, wrist pronation*). In addition, a *rest* state was defined when it was assumed the subject was not volitionally performing any of the motions and the residual musculature was in a neutral position. Following a previously described protocol [32], the subject performed each movement starting from a resting position and moving towards the motion end-state within one second according to an auditory cue. He then held the end state for one second, moved back to *rest* within one second, and remained at *rest* for another second. Thus, there were four phases per motion performance: *rest*, transition from *rest* to motion end-state, motion end-state, and transition from motion end-state to *rest*. Each motion performance was repeated five times, resulting in 11 separate 20-second sequences. A short break was provided between the collection of each sequence when the subject switched to a new motion.

### Data acquisition with electromyography

All data were acquired from the subject’s transhumeral residual limb. Raw EMG signals were gathered at a 1 kHz sampling rate with eight bipolar anodized titanium electrodes (Infinite Biomedical Technologies, Baltimore, MD) embedded within a custom transhumeral socket. The electrodes were amplified and filtered with a 20 - 500 Hz digital bandpass and 60 Hz digital notch filters. The subject created a single dataset containing the same set of 11 motions and following the same temporal sequence described previously. The only difference was that visual cues, rather than auditory cues, were provided to prompt transitions between states.

### Data analysis

The EMG and SMG sensing modalities produce very different data and were subjected to different pre-processing steps. The EMG data were split into 100 ms windows at every 10 ms interval. Five features were extracted from each of the windows: mean absolute value, variance, slope sign change count, waveform length, and number of zero crossings. The SMG data are acquired as a stream of individual ultrasound image frames. These ultrasound frames have a very large number of pixels and thus a very high dimensionality (1400×1000). To reduce the number of features, we first downscaled the images to one-tenth the original size (140×100), and then utilized principal component analysis to further reduce the dimensionality. We retained every basis vector that explained greater than 0.01% of the variance in the data, leading to a total explained variance of approximately 97%. Once the data were processed, each datapoint (time window for EMG, ultrasound image for SMG) was assigned a phase label based on the metronome cues. Thus, the datapoints were labeled as *rest* during the first second of motion performance, *transition to the motion end-state* during the next second, *motion end-state* during the third second, and *transition from the motion end-state* during the fourth second. This pattern of labelling was repeated for all five motion repetitions in the 20-second sequence.

Although the preprocessing was different for the two sensing strategies, the classification analysis for both the SMG and EMG datasets was performed using a linear discriminant analysis classifier. The classifier was constrained to have equal prior probabilities for each of the 12 possible classes (11 motions and *rest*). Leave-one-out cross-validation was performed such that one of the 55 motion performances (11 motions x 5 trials) were selected as a test set while the remaining 54 motion performances were used for training. This process was repeated for each of the 55 motion performances and a list of predictions for all data was compiled. Based on the phase labels, we then created confusion matrices and calculated the percent of correct predictions for each of the 4 phases, as well as the overall dataset.

For this work, we considered two different methods for training the classifier. Since we are primarily interested in evaluating the extent to which we can enable basic motion classification in individuals with transhumeral limb loss, we first trained the classifier using only data labelled as belonging to the *rest* and motion end-state phases where the user’s performance is most stable. However, we are also interested in understanding the extent to which we can facilitate an intuitive control paradigm that mimics the natural control employed with intact limbs. Our previous work has shown that SMG may eventually enable proportional positional control of prostheses [32] that allows users to rely on their innate proprioceptive sensing [34] to drive their device in a way that is directly congruent with their level of muscle contraction. Therefore, evaluating whether the classifier can accurately detect partially completed grasps is a necessary step towards demonstrating the feasibility of achieving proportional positional control. Being able to successfully detect partially completed grasps would also permit earlier classification of hand gestures than if the user was required to fully complete the grasp, thus increasing the responsiveness of the prosthetic device. As such, we also trained the classifier using data from the transition phases when grasps are partially completed, in addition to the *rest* and motion end-state phases.

Although the subject received cues to transition between each phase in the data acquisition sequence, it is likely that there were some delays due to reaction time following each cue. For example, they may have continued to remain at *rest* for a brief time even after receiving the cue to begin transitioning to the motion end-state. Thus, it is difficult to know whether a *rest* prediction during a transition phase is truly incorrect (i.e., misclassification of a motion state as *rest*) or correct (i.e., the subject was at *rest* and the classifier predicted *rest*). Because of this ambiguity, the classification performance is reported under both assumptions. The same classifier predictions were used to compute the classification accuracy in both scenarios. The only difference was that prediction of any motion during a transition phase as *rest* was counted as a failure when *rest* was considered incorrect, but was counted as a success when *rest* was considered correct. Together, the two scenarios provide both a conservative (*rest* considered incorrect) and best-case (*rest* considered correct) estimate of classification accuracy.

## Results

### Training with steady state phases only

Visual inspection of the predicted classifications throughout the entire data acquisition period indicates that the classifier usually predicted motion classes correctly for both modalities (Fig 1). However, the overall classification accuracy for all 11 motion classes plus *rest* was higher for SMG than EMG (Fig 2). This was true regardless of whether *rest* predictions during transitions were considered correct (EMG: 80.79%, SMG: 97.90%) or incorrect (EMG: 61.52%, SMG: 72.93%). Similarly, classification accuracy during the motion end-state phases (Fig 3) was higher for SMG (EMG: 75.08%, SMG: 94.04%). Classification accuracy during the *rest* phases was roughly equivalent between the two modalities (EMG: 96.25%, SMG: 98.34%).

**Fig 1.**
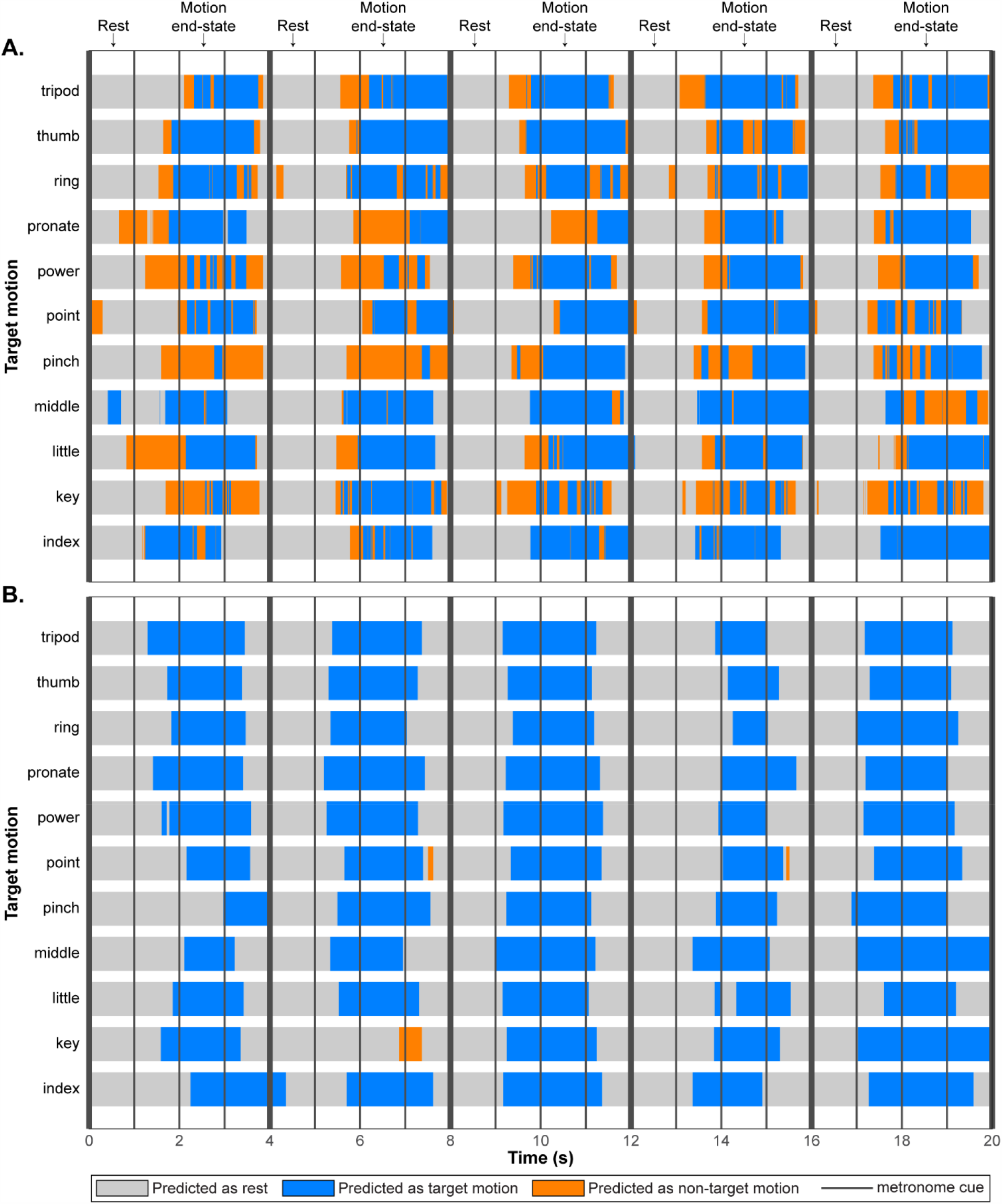
Motion class predictions over time. The temporal progression of motion class predictions is shown for all EMG time windows (**A**) and SMG image frames (**B**). Vertical lines represent metronome cues prompting a transition between phases. Each rest and motion end-state phase was followed by a one-second transition period during which the subject transitioned to the next phase. Rest predictions are indicated in grey, target motion predictions are indicated in blue, and predictions of any of the other 10 motions are indicated in orange.

**Fig 2.**
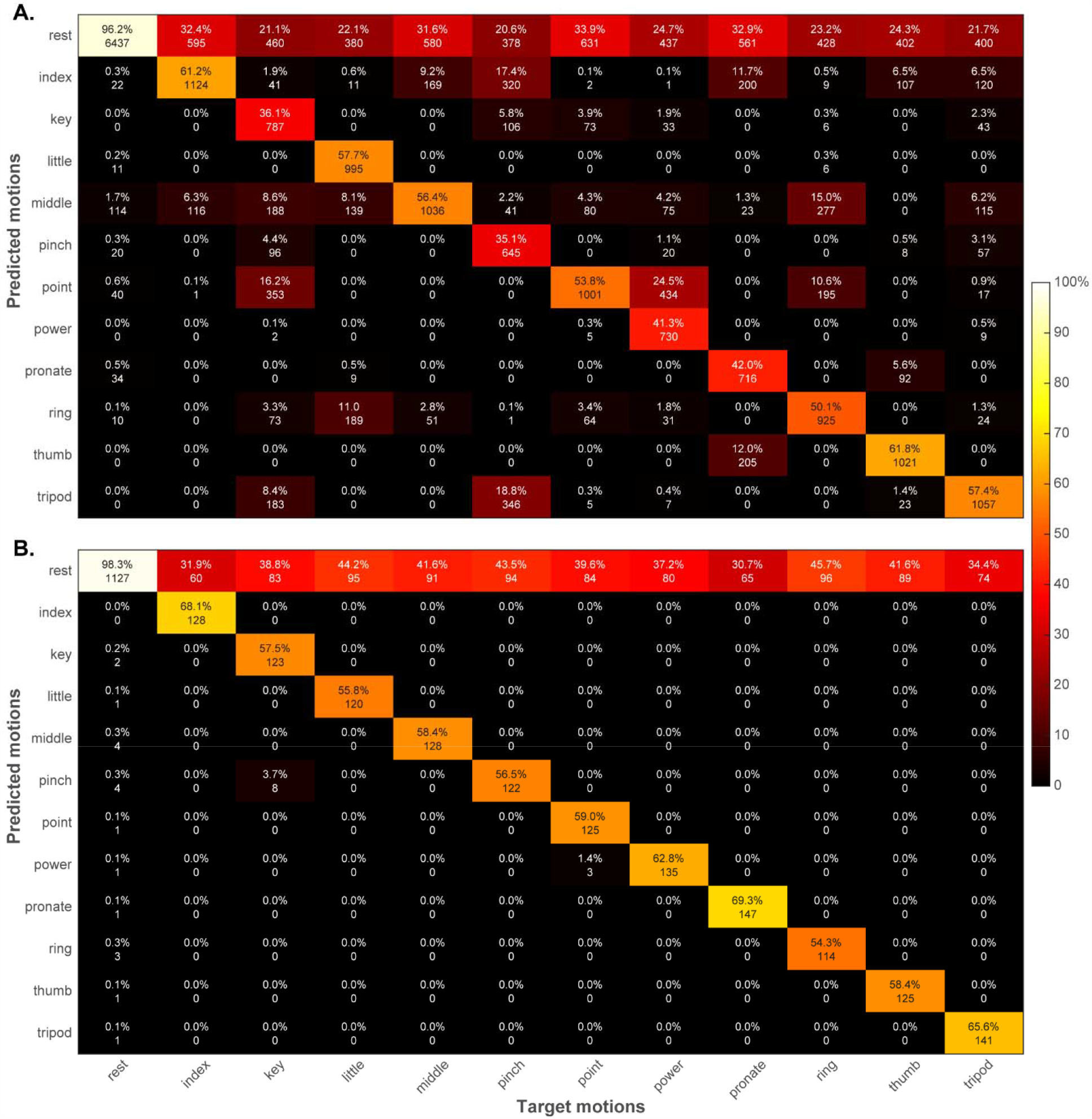
Overall confusion matrices. Prediction accuracies are shown for EMG (**A**) and SMG (**B**). The integer values in each cell represent the total number of EMG time windows or SMG image frames that were classified.

**Fig 3.**
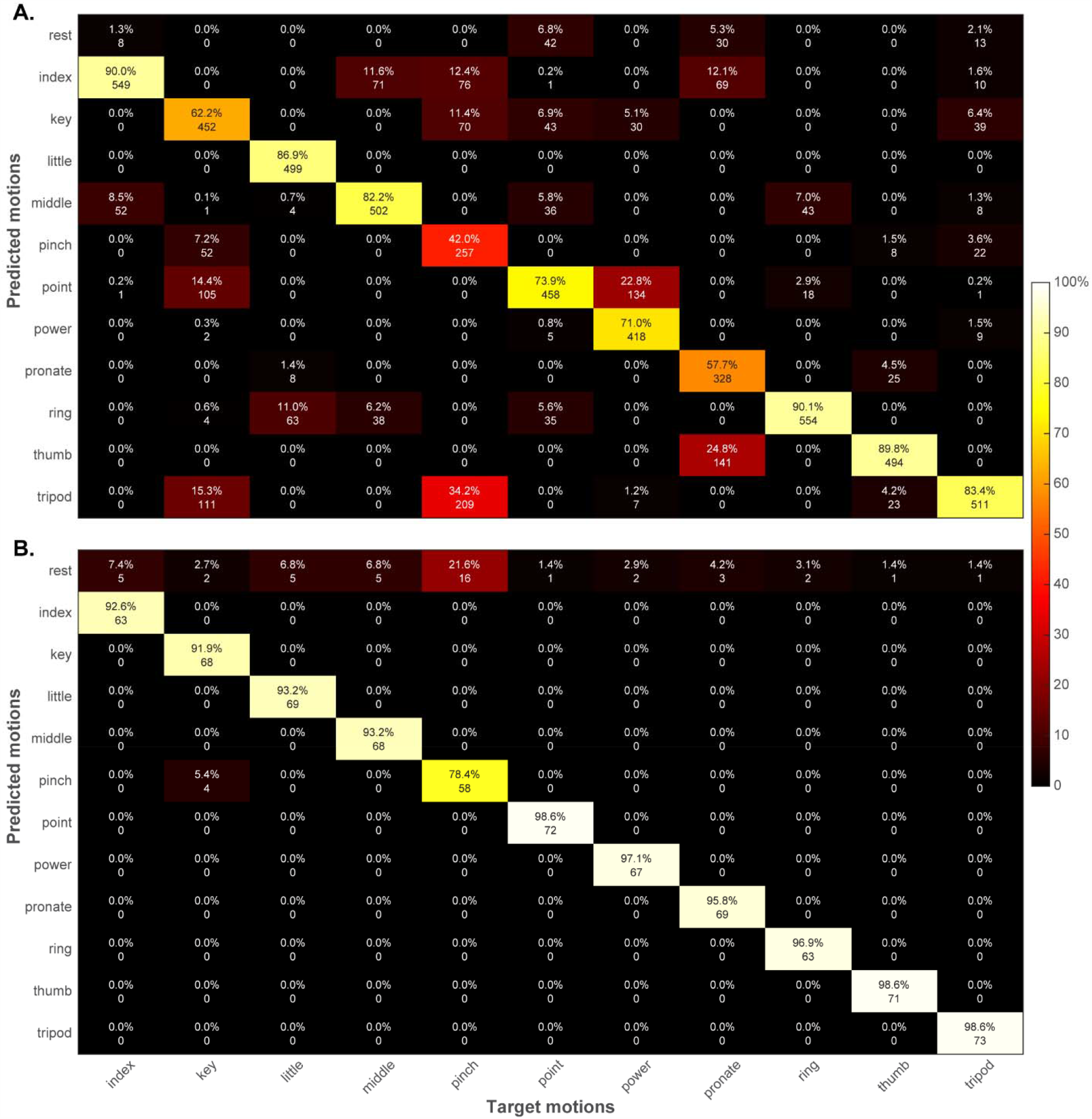
Confusion matrices for the motion end-state phases. Prediction accuracies are shown for EMG (**A**) and SMG (**B**). The integer values in each cell represent the total number of EMG time windows or SMG image frames that were classified.

During the transition periods from *rest* to a motion end-state, classification accuracy was higher for SMG (Fig 4) both when *rest* predictions were considered correct (EMG: 70.64%, SMG: 100.00%) and incorrect (EMG: 15.48%, SMG: 67.37%). During the transition periods from a motion end-state to *rest* (Fig 5), classification accuracy was higher for SMG only when *rest* predictions were considered correct (EMG: 81.16%, SMG: 99.09%). If *rest* predictions were considered incorrect, classification accuracy was higher for EMG (EMG: 59.31%, SMG: 18.93%). A summary of the classification accuracies for all phases is provided in Table 1.

**Table 1.** Classification accuracies when partially completed grasps were excluded from training.

| **Data acquisition phase** | **EMG** | **SMG** |
| --- | --- | --- |
| *Rest* | 96.25% | 98.34% |
| **Motion end-state** | 75.08% | 94.04% |
| *Rest predictions during transitions considered correct* |  |  |
| **Transition from *rest* to motion end-state** | 70.64% | 100.00% |
| **Transition from motion end-state to *rest*** | 81.16% | 99.09% |
| **All phases** | 80.79% | 97.90% |
| *Rest predictions during transitions considered incorrect* |  |  |
| **Transition from *rest* to motion end-state** | 15.48% | 67.27% |
| **Transition from motion end-state to *rest*** | 59.31% | 18.93% |
| **All phases** | 61.52% | 72.93% |

**Fig 4.**
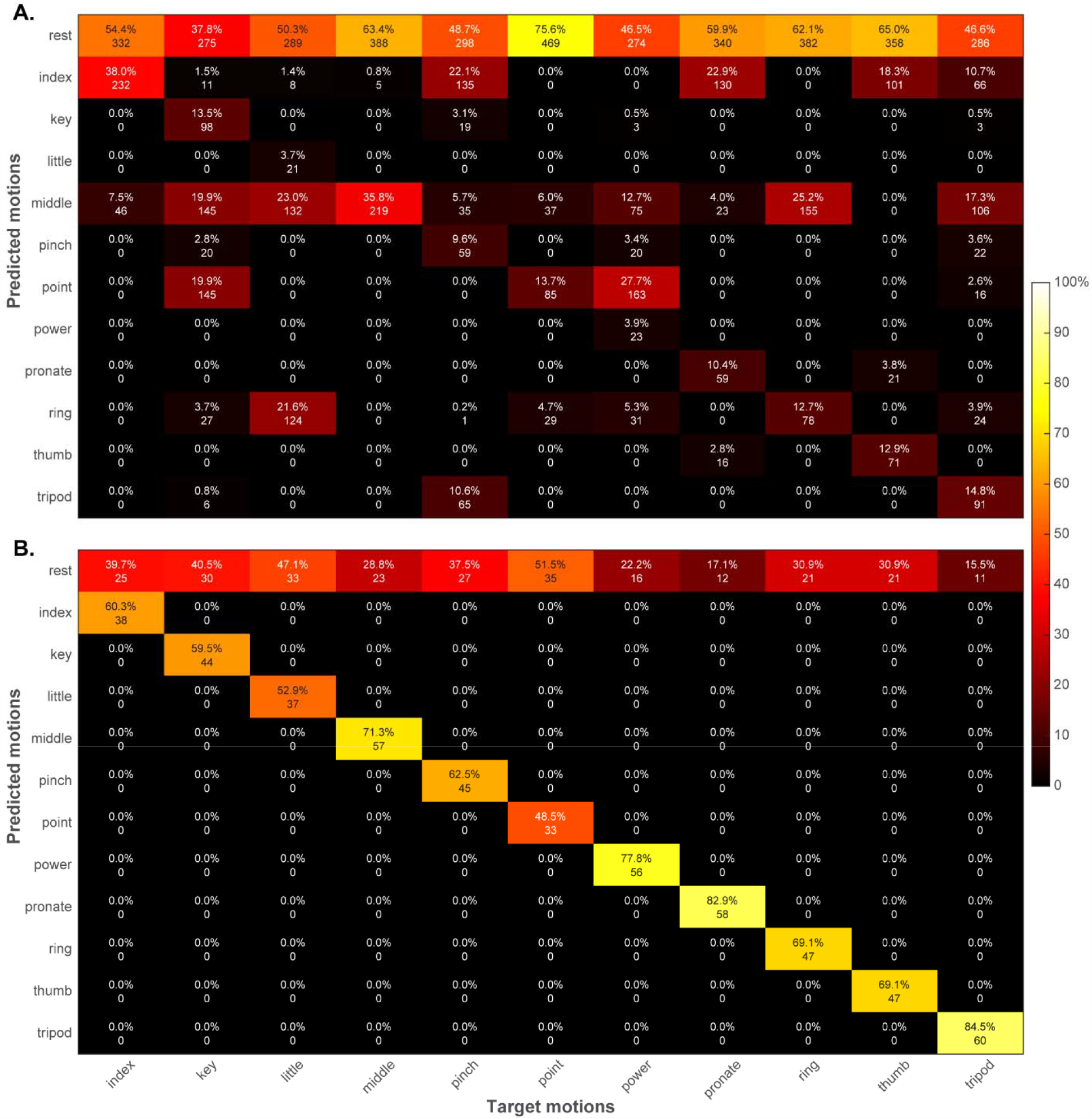
Confusion matrices for the transitions from rest to motion. Prediction accuracies are shown for EMG (**A**) and SMG (**B**). The integer values in each cell represent the total number of EMG time windows or SMG image frames that were classified.

**Fig 5.**
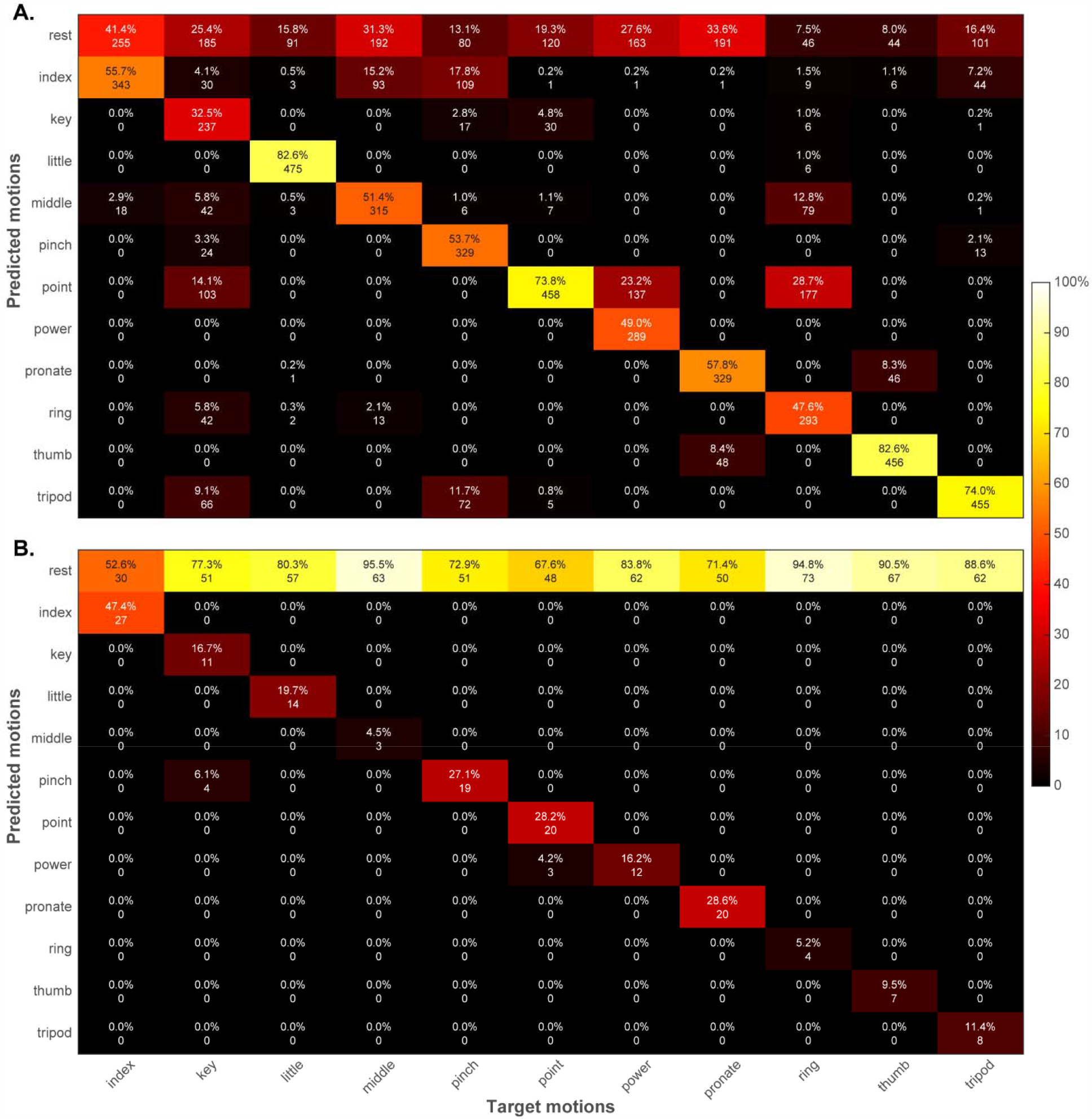
Confusion matrices for the transitions from motion to rest. Prediction accuracies are shown for EMG (**A**) and SMG (**B**). The integer values in each cell represent the total number of EMG time windows or SMG image frames that were classified.

### Training with steady state and transition phases

When partially completed grasps were included in the training, the classifier usually predicted motion classes correctly (Fig 6) but with higher overall accuracy for SMG than EMG (Fig 7). The accuracy was higher for SMG both when *rest* predictions during transitions were considered correct (EMG: 76.44%, SMG: 87.26%) and incorrect (EMG: 62.38%, SMG: 79.63%). Classification accuracy was also higher for SMG during the motion end-state phases (Fig 8, EMG: 77.53%, SMG: 97.34%) but was higher for EMG during the *rest* phases (EMG: 78.29%, SMG: 63.44%).

**Fig 6.**
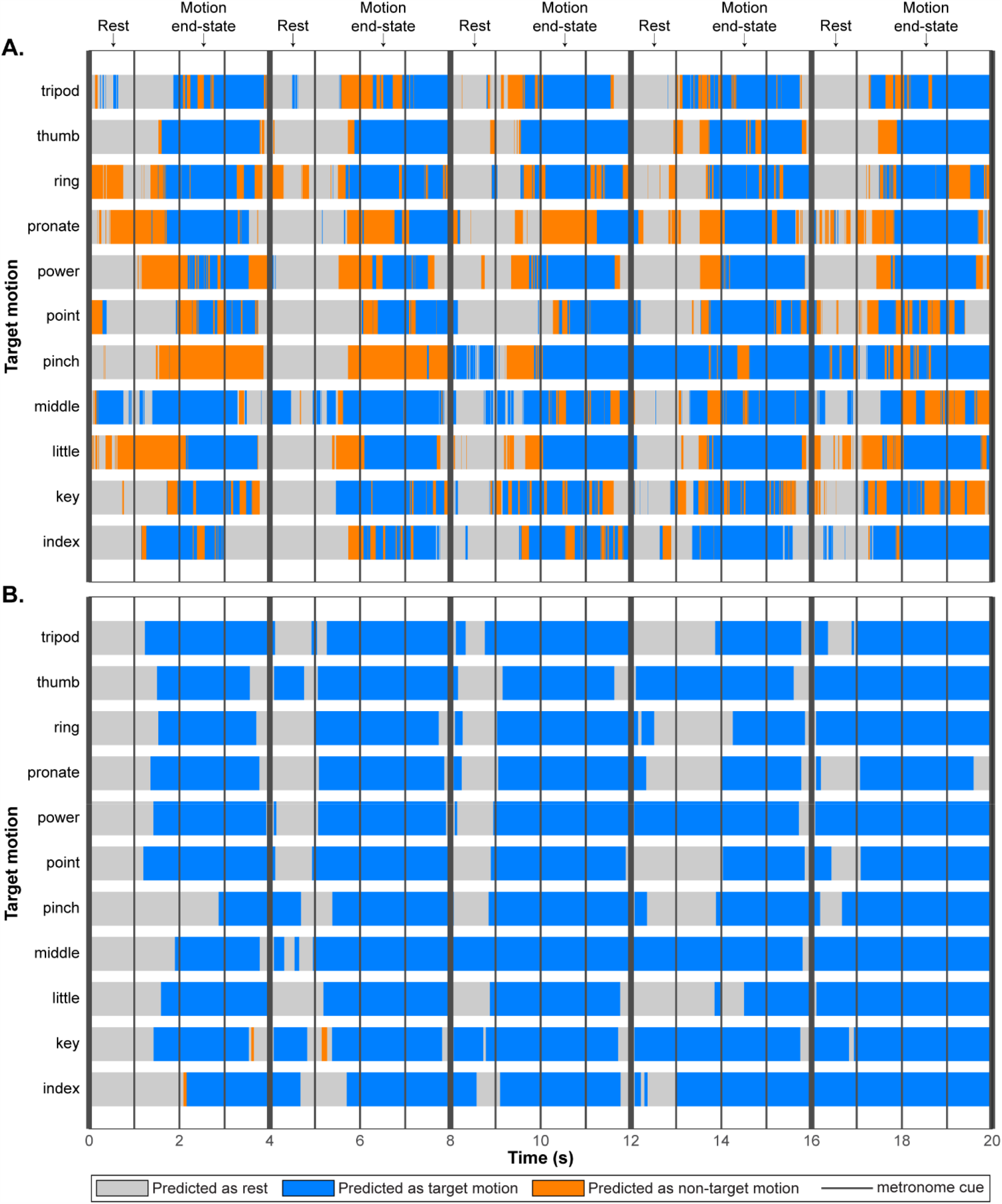
Motion class predictions over time. The temporal progression of motion class predictions is shown for all EMG time windows (**A**) and SMG image frames (**B**). Vertical lines represent metronome cues prompting a transition between phases. Each rest and motion end-state phase was followed by a one-second transition period during which the subject transitioned to the next phase. Rest predictions are indicated in grey, target motion predictions are indicated in blue, and predictions of any of the other 10 motions are indicated in orange.

**Fig 7.**
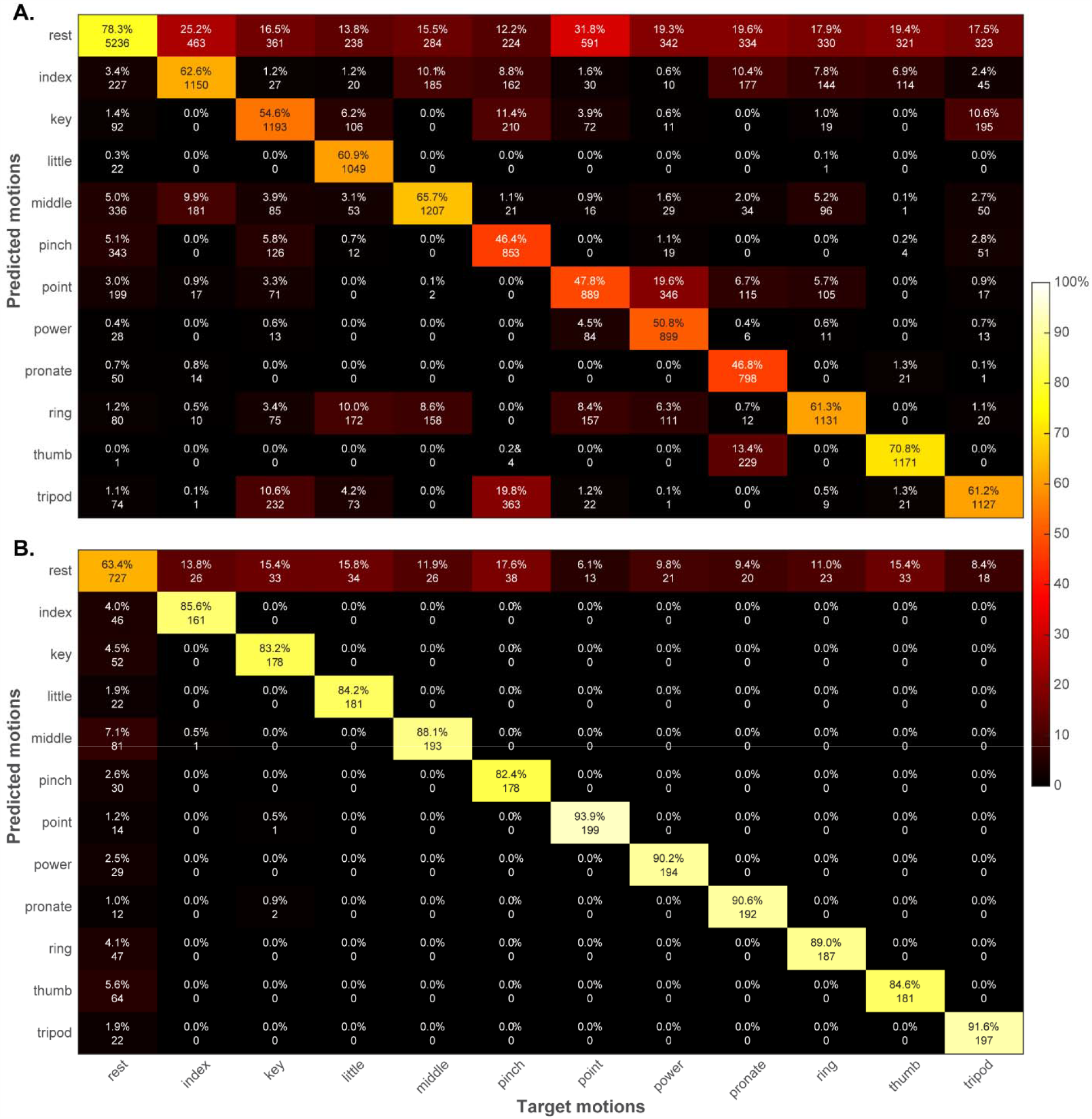
Overall confusion matrices. Prediction accuracies are shown for EMG (**A**) and SMG (**B**). The integer values in each cell represent the total number of EMG time windows or SMG image frames that were classified.

**Fig 8.**
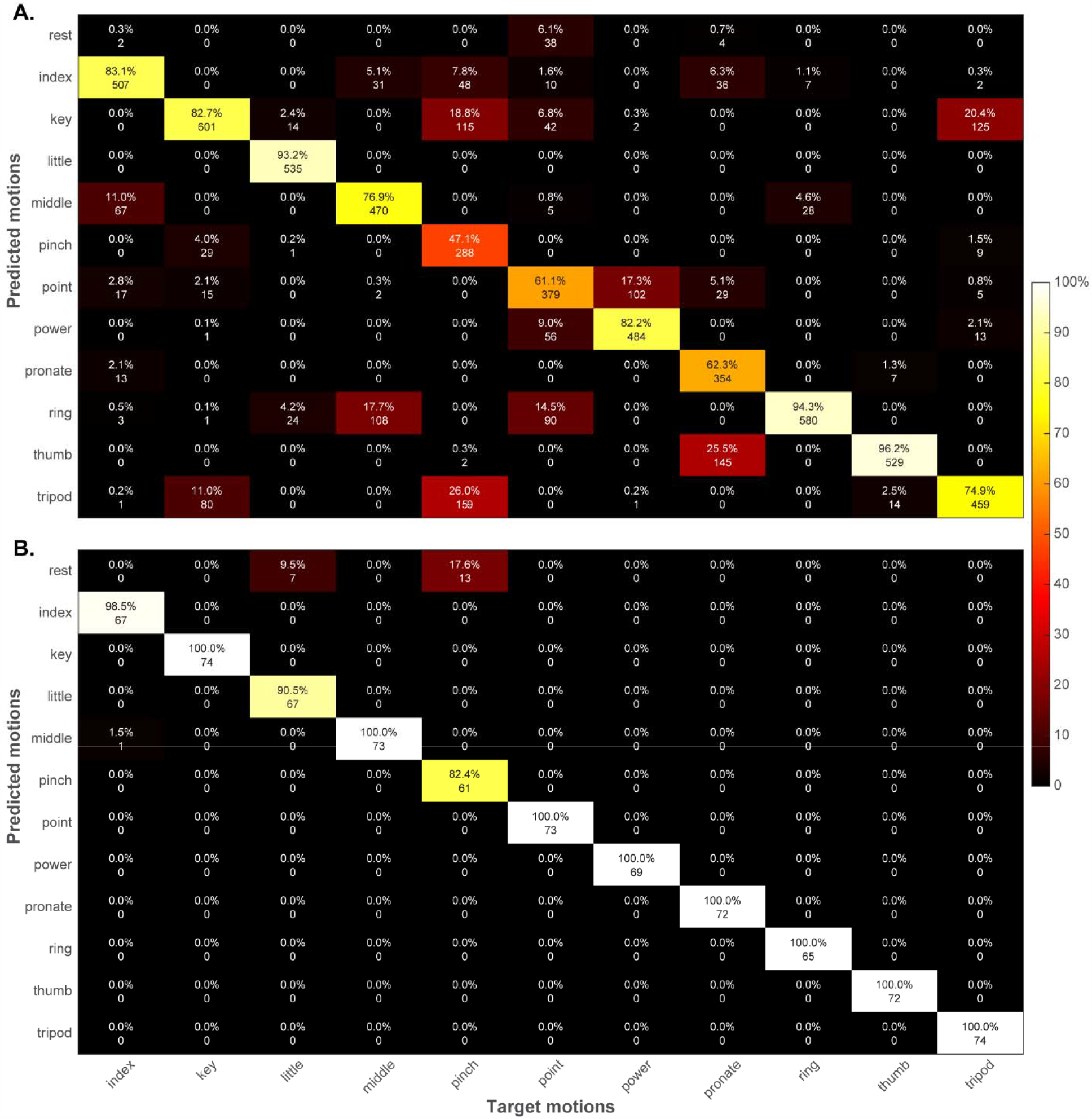
Confusion matrices for the motion end-state phases. Prediction accuracies are shown for EMG (**A**) and SMG (**B**). The integer values in each cell represent the total number of EMG time windows or SMG image frames that were classified.

During transitions from *rest* to a motion end-state (Fig 9), classification accuracy was higher for SMG if *rest* predictions were considered correct (EMG: 69.49%, SMG: 99.74%) or incorrect (EMG: 27.50%, SMG: 85.05%). Classification accuracy was also higher for SMG during transitions from a motion end-state to *rest* (Fig 10). This was true regardless of whether *rest* predictions were considered correct (EMG: 80.46%, SMG: 99.87%) or incorrect (EMG: 66.19%, SMG: 80.16%). A summary of the classification accuracies for all phases is provided in Table 2.

**Table 2.** Classification accuracies when partially completed grasps were included in training.

| Data acquisition phase | EMG | SMG |
| --- | --- | --- |
| *Rest* | 78.29% | 63.44% |
| **Motion end-state** | 77.53% | 97.34% |
| *Rest predictions during transitions considered correct* |  |  |
| Transition from *rest* to motion end-state | 69.49% | 99.74% |
| Transition from motion end-state to *rest* | 80.46% | 99.87% |
| **All phases** | 76.44% | 87.26% |
| *Rest predictions during transitions considered incorrect* |  |  |
| Transition from *rest* to motion end-state | 27.50% | 85.05% |
| Transition from motion end-state to *rest* | 66.19% | 80.16% |
| **All phases** | 62.38% | 79.63% |

**Fig 9.**
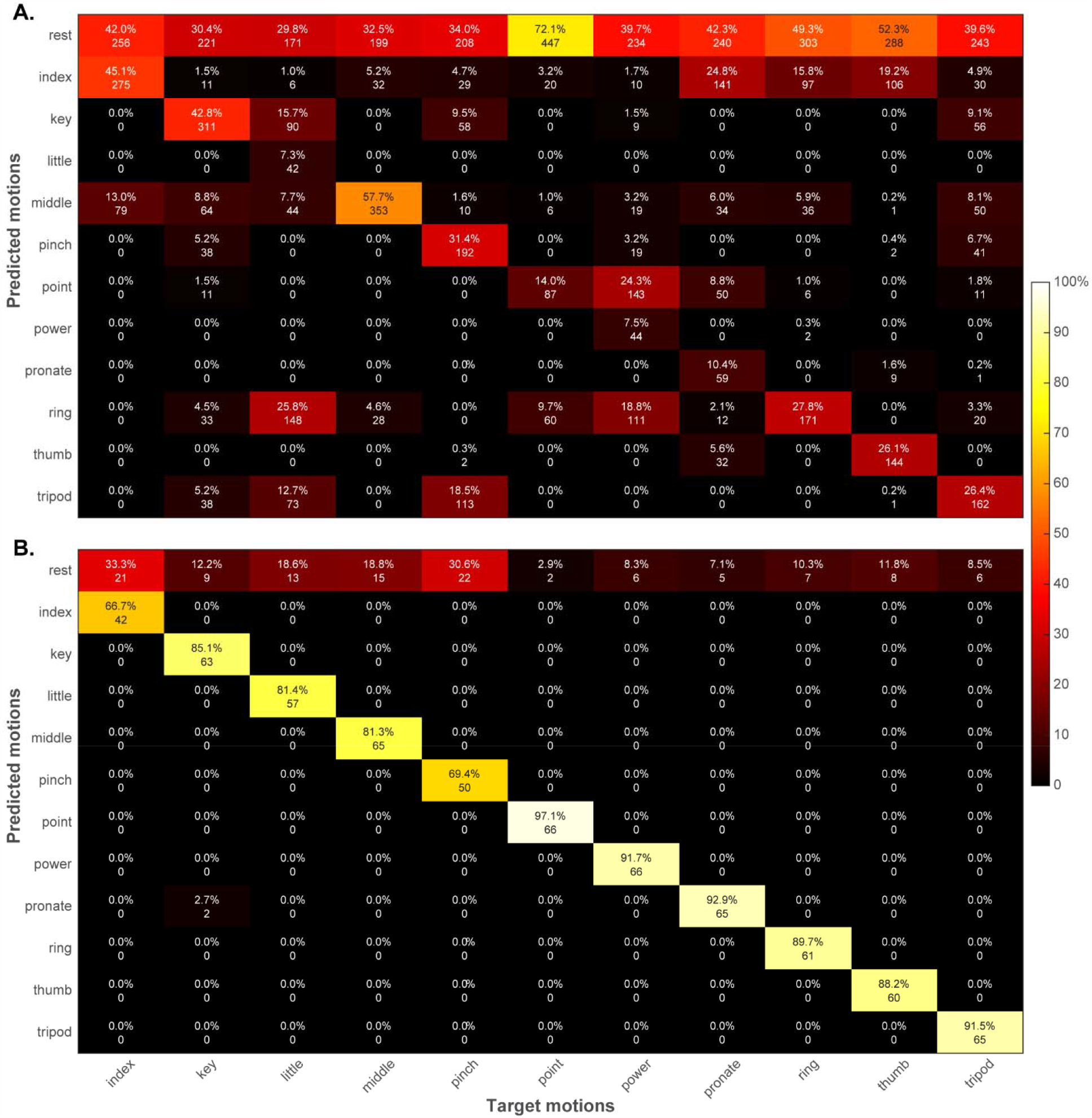
Confusion matrices for the transitions from rest to motion. Prediction accuracies are shown for EMG (**A**) and SMG (**B**). The integer values in each cell represent the total number of EMG time windows or SMG image frames that were classified.

**Fig 10.**
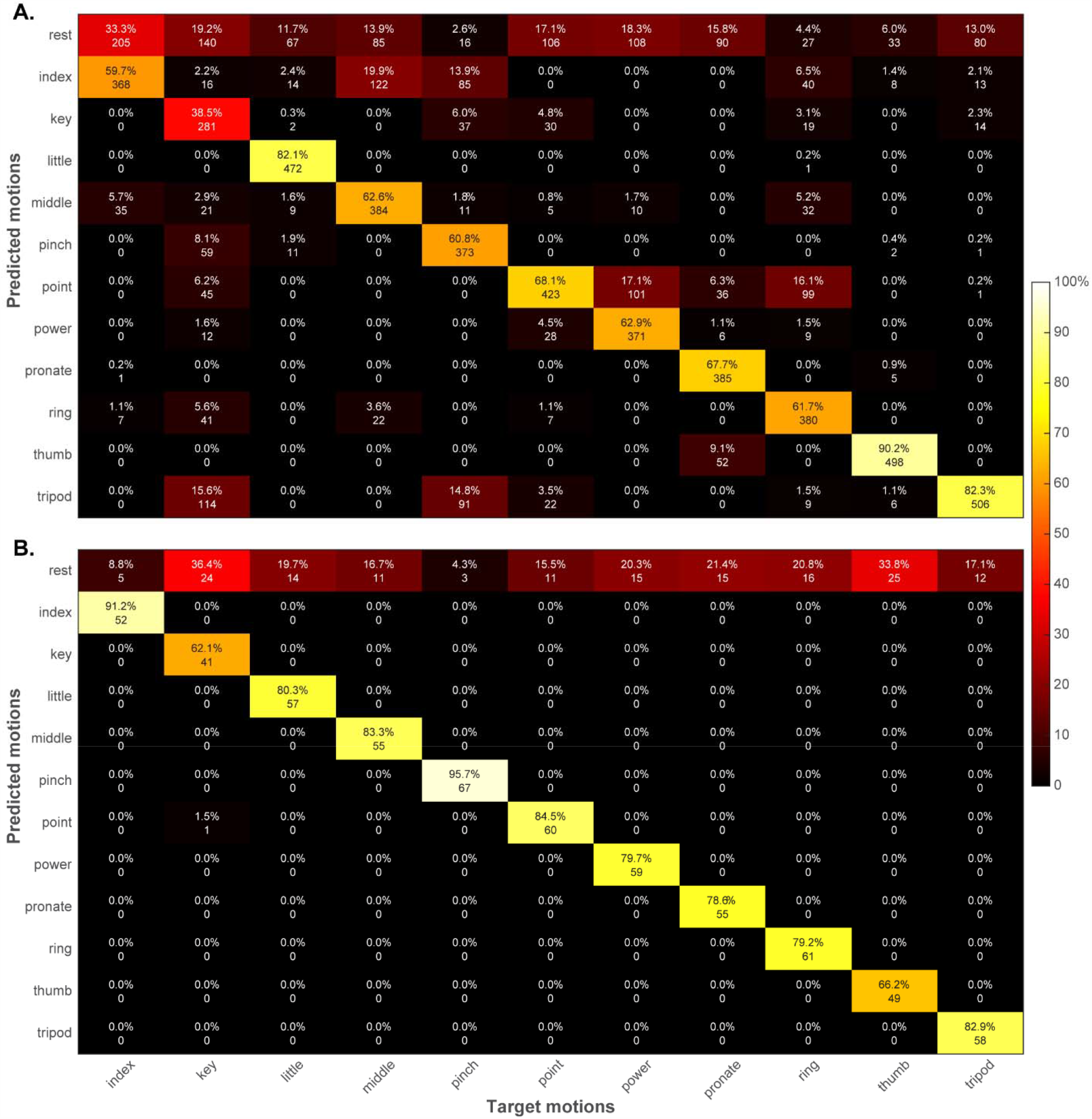
Confusion matrices for the transitions from motion to rest. Prediction accuracies are shown for EMG (**A**) and SMG (**B**). The integer values in each cell represent the total number of EMG time windows or SMG image frames that were classified.

## Discussion

This work provides the first documentation of successful motion classification in an individual with transhumeral limb loss using SMG. Notably, the subject was able to accomplish both individual finger movements and complex grasps. Recognizing finger motions with EMG is challenging since the muscles responsible for finger movement are largely contained within the intermediate and deep compartments of the forearm, meaning that the electrical activity is attenuated by the overlying tissue [20]. SMG appears to be more well-suited for classifying individual digit motion given its ability to detect deformation in both superficial and deep muscles [27,35,36]. Nonetheless, classifying individual finger motion could be problematic in individuals with transhumeral limb loss given the complete absence of forearm musculature.

Our subject’s ability to successfully classify both individual finger movements and complex grasps using SMG may be a consequence of procedures used during his amputation surgery. The surgeons removed necrotic tissue and deinnervated some muscles in the residual limb. We hypothesize that these muscles may have spontaneously reinnervated during recovery, although this hypothesis could not be directly tested and only indirect inference is possible. The surgery was not performed with the expectation that reinnervation would occur. Therefore, if our hypothesis is true, this case could be evidence of spontaneously-occurring muscle reinnervation [37]. As a planned surgical procedure, targeted muscle reinnervation has shown great promise for restoring wrist, hand, and finger control to patients with proximal limb loss (e.g., [13]). During surgery, severed nerves that previously controlled arm and hand function are transferred to muscles in the residual limb that no longer serve a biomechanical purpose. Thus, an attempt to move the missing arm or hand will result in contraction of the reinnervated muscles. The individual can then generate a wider variety of distinct muscle contractions than if the muscles had not been transferred. Since our subject shows similar diversity in his residual limb movement when performing the 11 gestures used in this study, we hypothesize that spontaneous reinnervation may have occurred.

If spontaneous reinnervation was indeed responsible for our subject’s robust classification performance, it is possible that other individuals who have undergone more traditional transhumeral amputations would be unable to achieve similar levels of success. A larger group of individuals with transhumeral limb loss must be studied to determine whether SMG can be a viable control strategy for this population. It would also be interesting to consider the use of SMG as an alternative to EMG pattern recognition in those who have formally undergone targeted muscle reinnervation.

A secondary aim of this work was to directly compare motion classification using EMG and SMG in order to understand the relative merits of each modality. In general, the motion classification accuracy was higher for SMG than EMG. This was true regardless of whether partially completed grasps were excluded or included during training. There were only two exceptions. First, the classification accuracy was higher for EMG during transitions from a motion end-state to *rest* if *rest* predictions were assumed to be incorrect and training was performed without partially completed grasps. Second, the classification accuracy was higher for EMG during *rest* if training was performed with partially completed grasps.

Within each modality, the effect of including partially completed grasps was inconsistent between phases. During motion end-state phases, classification accuracy was similar within each modality whether or not partially completed grasps were included during training. However, including partially completed grasps during training reduced classification accuracy for *rest* phases, especially for SMG. Classification accuracies during transitions were similar within each modality regardless of whether partially completed grasps were included during training, but only if *rest* was considered a correct prediction. If *rest* was considered incorrect, including partially completed grasps increased classification accuracy within each modality.

Taken together, these findings indicate that including partially completed grasps during training does not necessarily improve classification accuracy and also comes at the expense of deterioration in *rest* prediction accuracy. This reduction in accuracy might occur because low-level muscle activity can be classified as movement if partially completed grasps are included in training. If only *rest* and motion end-states were included in training, then low-level muscle activity would more likely be classified as *rest* until it became pronounced enough to resemble a motion end-state. This issue could be exacerbated by the high spatial specificity of SMG, which enables the detection of minute muscular deformations. Additionally, muscular *rest* states are not constant since the muscles retain some residual deformation even after a contraction is completed. However, inclusion of partial grasps during training did not reduce prediction accuracy during transitions.

Although the reduction in *rest* prediction accuracy may appear undesirable, we do not believe it will be overly problematic. We intend to use SMG to implement proportional positional control strategies [32], in which the finger positions on a prosthetic hand are mapped proportionally to the amount of muscle deformation in a user’s residual limb. In this paradigm, misclassifying some *rest* states as motion may be tolerable since each motion prediction would be accompanied by a predicted motion completion level. Thus, misclassification of *rest* as a partially completed motion could be considered a more reasonable trade-off than misclassifying *rest* as a fully completed motion, which would occur if proportional control is not enabled. However, if this trade-off proves to be intolerable from a user’s perspective, other approaches may be considered to improve the stability of the *rest* predictions while retaining the benefits of SMG-based control (namely, robust classification of motion end-states and the ability to enable proportional positional control). One possibility could be the implementation of a hybrid control system in which EMG signals serve as the input for a binary classifier to predict *rest* or motion. Once a deviation from *rest* is detected, then SMG could be used for classifying the intended grasp and completion level.

One limitation that should be acknowledged about this study is that only a single participant was involved, so the findings should not be generalized without additional study in a larger population of individuals with transhumeral limb loss. Additionally, the modality used for prompting the subject’s transitions between motion and *rest* states was inconsistent (i.e., auditory cues for SMG, visual cues for EMG). The visual cues were used as a matter of convenience, as the subject collected the EMG data from himself. Reaction times for auditory cues are typically shorter than those for visual cues [38,39], with a commonly-reported difference of approximately 40 ms [40]. We do not believe a difference of this magnitude would have any bearing on the results. Furthermore, both the EMG and SMG data were collected under ideal settings with no external sources of noise that would likely arise during real-world prosthesis use (e.g., changes in skin impedance from perspiration, shifts in the relative positions of the residual limb and sensors due to arm movement or socket loading). These problems are known to occur with our subject’s clinically prescribed EMG prosthesis, so he typically has only six motion classes enabled. Predicting 11 motion classes would be extremely difficult for him under normal circumstances. Although we are currently undertaking work to formally assess the use of SMG to control prostheses in real-world settings, we anticipate that similar issues may arise that would reduce the number of achievable motion classes. Additionally, it should be reiterated that the SMG data were obtained using a clinical ultrasound system with an array transducer. In the future, we plan to use single-element transducers with low-power electronics that could be integrated into a standalone prosthesis socket. Although our prior work suggests that classification accuracy is not reduced with a sparse sensing strategy [41], this has not yet been confirmed in individuals with limb loss. Finally, the sampling rate for EMG was considerably higher than for SMG, so more frames of EMG data were available to train the classifier. However, the use of a smaller training data set for SMG did not appear to have a detrimental effect, as the classification accuracy was higher for SMG than EMG in most circumstances.

## Conclusion

This exploratory study demonstrated that prediction of 11 hand and finger motions was possible for an individual with transhumeral limb loss using SMG. Despite the individual’s lack of a forearm, he was able to generate distinctive muscle activation patterns in his upper arm that could be successfully detected and classified using both SMG and EMG. The classification accuracy was typically higher for SMG than EMG, regardless of the method used to train the classifier. These findings suggest that SMG may eventually be a viable prosthesis control strategy, even for individuals with transhumeral limb loss.

## Data Availability

All data are available from the authors upon reasonable request.

## Acknowledgements

The authors would like to thank Dr. Jaimie Shores for sharing his expertise regarding amputation procedures and targeted muscle reinnervation.

## Supporting Information

**S1 Dataset. Full datasets collected using SMG and EMG**.

